# Ketamine-assisted psychotherapy provides lasting and effective results in the treatment of depression, anxiety and post traumatic stress disorder at 3 and 6 months: Findings from a large single-arm retrospective effectiveness trial

**DOI:** 10.1101/2023.01.11.23284248

**Authors:** Ryan Yermus, Michael Verbora, Sidney Kennedy, Robert McMaster, Sarah Kratina, Elizabeth Wolfson, Ben Medrano, Nathan Bryson, Nabid Zaer, John Bottos, Varun Setlur, Chris Lo

**Affiliations:** Field Trip Health, Toronto, Ontario; Family Medicine, Faculty of Health Sciences, McMaster University, Hamilton, Ontario; Department of Psychiatry, Temerty Faculty of Medicine, University of Toronto, Toronto, Ontario; Institute of Health Policy, Management and Evaluation, Dalla Lana School of Public Health, University of Toronto, Toronto, Ontario; Social and Behavioural Health Sciences Division, Dalla Lana School of Public Health, University of Toronto, Toronto, Ontario; Psychology, School of Social and Health Sciences, James Cook University, Singapore; Field Trip Health NY & DC, New York, NY & Washington, District of Colombia; Nue Life Medical Group, Delaware; Reunion Neuroscience, Toronto, Ontario; School of Biological Sciences and Applied Chemistry, Seneca College, Toronto, Ontario

**Keywords:** Anxiety, Depression, Ketamine, Psychedelic, Psychotherapy, Post Traumatic Stress Disorder

## Abstract

**IMPORTANCE:** Ketamine-Assisted Psychotherapy (KAP) is an emerging treatment option to alleviate treatment resistant affective disorders, but its long term effectiveness remains unclear.

**OBJECTIVE:** To examine the treatment effects of KAP on anxiety, depression, and post traumatic stress disorder (PTSD) at 1, 3, and 6 months post treatment.

**DESIGN, SETTING, AND PARTICIPANTS:** This retrospective single-arm effectiveness trial included self-reported outcomes from 1806 adults with a history of depression, anxiety, or PTSD who had not responded to prior treatment interventions and received KAP administered across 11 Field Trip Health clinics in North America between March 13, 2020 and June 16, 2022.

**INTERVENTION:** KAP consisting of 4-6 guided ketamine sessions (administered via intramuscular injection or sublingual lozenge) with psychotherapy-only visits after doses 1 and 2 and then after every 2 subsequent doses. Mean number of doses administered was 4, SD=3, and mean number of psychotherapy sessions was 3, SD=2.

**MAIN OUTCOMES AND MEASURES:** Primary outcomes were changes in depression, anxiety, and PTSD at 3 months relative to baseline, assessed respectively using the 9-item Patient Health Questionnaire (PHQ-9), the 7-item Generalized Anxiety Disorder measure (GAD-7), and the 6-item PTSD Checklist (PCL-6). Secondary outcomes were changes at 1 and 6 months relative to baseline.

**RESULTS:** Large treatment effects were detected at 3 months (d’s=0.75-0.86) that were sustained at 6 months (d’s=0.61-0.73). Case reductions (identified based on cut-off values) ranged from 39-41% at 3 months and 29-37% at 6 months. 50-75% reported a minimal clinically important difference at 3 months and 48-70% at 6 months.

**CONCLUSIONS AND RELEVANCE:** KAP produced sustained reductions in anxiety, depression, and PTSD, with symptom improvement lasting well beyond the duration of dosing sessions. These effects extended to as much as 5 months after the last KAP session. Given the growing mental health care crises and the need for effective therapies and models of care, especially for intractable psychiatric mood related disorders, these data would support the consideration of KAP as a viable alternative. Further prospective clinical research should be undertaken to provide further evidence on the safety and effectiveness of ketamine within a psychotherapeutic context.

**TRIAL REGISTRATION:** Clinicaltrials.gov Identifier NCT05604794

**Key Points:** 

**Question:** What are the lasting effects of Ketamine-Assisted Psychotherapy on psychological distress?

**Findings:** In this retrospective single-arm effectiveness trial that included 1806 adults, there were large effect sizes at 3 months on depression, anxiety, and post traumatic stress (d’s=0.75-0.86) that were sustained at 6 months.

**Meaning:** These findings suggest that Ketamine-Assisted Psychotherapy is an effective treatment option with substantial clinical benefits detected up to half a year.

## Introduction

Over the past two decades, ketamine has demonstrated the potential to produce rapid and sustained antidepressant effects and therapeutic outcomes for several psychiatric conditions.^1^ Most studies to date have looked at the administration of ketamine via intravenous (IV) administration^2^ however, ketamine’s application has evolved to include psychotherapeutic practices to reduce its unwanted dissociative effects while bolstering positive antidepressant, anxiolytic, anti-stress, and other positive mood effects.^3^ Ketamine-Assisted Psychotherapy (KAP) has borrowed from earlier research on psychedelic substances such as lysergic acid diethylamide (LSD) to facilitate deep and rapid introspective work within an organized psychotherapeutic framework.^4^

Currently, ketamine is the only legal psychedelic medicine to treat persistent and severe psychological distress in North America, making it well-suited for widespread psychotherapeutic implementation. Preliminary clinical trial data are increasingly supportive of the safety and efficacy of KAP to alleviate anxiety, depression, and post traumatic stress disorder (PTSD),^5,6^ with more trials underway worldwide. In a recent study of patients with moderate to severe depression or anxiety, ketamine-assisted therapy was found to have immediate effects on depression and anxiety; outcomes showed persistent efficacy to 2 and 4 weeks in most treated subjects.^7^ However, the impact on post-treatment quality of life is understudied, and there remains a need to clarify the long term effectiveness of KAP for a variety of mood related disorders.

In this study, we conducted a retrospective analysis of outcomes from patients who were treated with KAP at Field Trip Health centres across North America and agreed to participate in an open label evaluation of outcomes up to 6 months after the intervention. Ketamine was administered via intramuscular injection or sublingual lozenge. Primary outcomes were changes in depression, anxiety, and post traumatic stress at 3 months from baseline. Secondary outcomes were changes at 1 and 6 months from baseline.

## Methods

### Ethics and Design

Ethics approval for this study was granted by Veritas Independent Review Board (#2022-3067-11240-5). The study was a retrospective single-arm effectiveness trial of KAP involving chart review of patient self-reported mental health outcomes assessed at baseline, 1, 3, and 6 months.

### Participants and Procedure

Data was collected from clients treated across 11 Field Trip Health clinics in North America: 1) Toronto, ON, Canada; 2) Vancouver, BC, Canada; 3) Fredericton, NB, Canada; 4) New York City, NY, USA; 5) Atlanta, GA, USA; 6) Chicago, IL, USA; 7) Houston, TX, USA; 8) Seattle, WA, USA; 9) Santa Monica, CA, USA; 10) San Diego, CA, USA; 11) Washington, DC, USA; between March 13, 2020, and June 16, 2022. Participants included in the analysis had a documented history of depression or anxiety that showed lack of adequate response to previous treatment(s) or presented with PTSD as assessed by the Structured Clinical Interview for DSM 5 (SCID 5).^8^ Patient assessments were performed using a proprietary digital platform (PortalTM) through the Electronic Medical Records (EMR) system at 1, 3, and 6 months. The completion of measures was voluntary after the collection of signed written informed consent.

### Treatment

Prospective clients were either self-referred in the United States or referred for treatment by a health care provider to one of the 3 Canadian clinic locations. All clients were assessed by a psychiatrist or psychiatric nurse practitioner to determine the appropriateness for treatment. Inclusion criteria included signed written informed consent; being over the age of 18; and having a documented, prior diagnosis by a psychiatrist of one or more of Major Depressive Disorder (MDD), Bipolar Depression, Generalized Anxiety Disorder, Obsessive Compulsive Disorder (OCD), Eating Disorder, or a significant history of trauma and/or a formal diagnosis of PTSD as per the DSM 5.

Exclusion criteria included pregnant women and nursing mothers, although Postpartum Depression (PPD) was considered on a case-by-case basis in consultation with the National Medical Director; a relative (not absolute) contraindication for individuals with a Body Mass Index (BMI) above 35; any individual who has met DSM 5 criteria for a Substance Use Disorder in the past 3 months and had been unable to exhibit a reduction in use; Psychosis or psychotic symptoms; Active Mania: Bipolar 1 (chronic non-disruptive hypomania is an exception at the discretion of the treatment team); Borderline Personality Disorder; uncontrolled medical disorders or physical conditions with negative interaction with ketamine; individuals with symptomatic acute brain injury within 90 days of serious injury; individuals diagnosed with moderate to severe sleep apnea; and individuals who were unable to identify a person or service to assure their safe transport to home post treatment.

Upon medical approval, clients met with a licensed therapist to discuss preparation for KAP and to initiate a therapeutic relationship. Dosing with ketamine was completed via intramuscular injection in the United States and Vancouver and via sublingual lozenge in Toronto and Fredericton. Clients were offered personalized treatment recommendations consisting of 4-6 guided ketamine sessions with psychotherapy-only integration visits after doses 1 and 2 and then after every 2 subsequent doses.

Initial dosing via intramuscular injection was of 25-35mg with the option to titrate to 50-70mg at visit 2 and up to 100mg for subsequent visits. The initial dose for lozenges was 200mg with the option to increase by 50-100mg per visit up to 500mg. Ketamine doses were not scheduled on consecutive days and could be interspersed by 1 week or more. Some participants completed additional sessions beyond dose 6. Integration sessions were based on motivational interviewing and behavioural activation, but therapists had the option to incorporate other modalities depending on their assessment of the client’s needs and goals.

Ketamine dosing was completed in Field Trip Health clinics using an approach consistent with psychedelic studies in a setting designed to be aesthetically and functionally conducive with a state of relaxation. Clients were dosed while seated in a comfortable reclining chair, wearing an eye shade and listening to curated music playlists. Therapists were present to support clients during dosing sessions while medical staff monitored heart rate, blood pressure, respiration rate and oxygen saturation throughout the session.

### Outcome Measures

Symptoms of depression were assessed by the 9-item Patient Health Questionnaire (PHQ-9).^9^ PHQ scores may range from 0-27 with higher scores indicating more severe depressive symptoms. A PHQ cut-off score of 15 has been validated to identify cases with at least moderately-severe depression.^9^ A change of 3 points has been considered a minimal clinically important difference (MCID).^10,11^

Symptoms of anxiety were assessed by the 7-item Generalized Anxiety Disorder measure (GAD-7).^12^ GAD scores may range from 0-21 with higher scores indicating more severe anxiety symptoms. A GAD cut-off score of 10 has been validated to identify cases with at least moderate anxiety.^12^ A change of 3 points has been considered an MCID.^10,11^

Symptoms of post traumatic stress were assessed using the 6-item PTSD Checklist (PCL-6).^13^ PCL scores may range from 6-30, with higher scores indicating more severe stress symptoms. A PCL cut-off score of 14 has been validated to identify cases of PTSD.^13^ A change of 5 points may be considered an MCID.^14^

### Statistical Analysis

We report descriptive statistics for the sample and analyze the extent of loss to follow-up. The primary analysis was by Intention to Treat (ITT). We used linear mixed modelling to fit growth curves describing the normative patient trajectory on each outcome.^15^ The main analysis involved fitting linear and curvilinear trends over time and estimating mean differences at each endpoint compared to the baseline. Cohen’s d was reported as a standardized measure of effect size for mean differences (d=0.2 is a small effect, 0.5 medium, and 0.8 large).

We reported secondary analyses on the effect of doses administered, controlling for age, gender, and site differences; case reductions in depression, anxiety, and PTSD based on cut-offs; and proportions of treatment responders based on MCIDs. As a sensitivity analysis, we used the Expectation-Maximization (EM) and Markov Chain Monte Carlo (MCMC) algorithms to multiply impute missing follow-up data.^16^ We simulated 1000 datasets per outcome measure, calculated the effects per imputation, and combined the findings across imputations to achieve the best estimates for comparison to the initial estimates.

## Results

### Descriptive Statistics

In total 1806 participants entered treatment (see Table 1 for sample characteristics). The mean age was 42 years, SD=12, and 52% of participants were female. Most individuals had a primary diagnosis of depression (24%), anxiety (28%), or PTSD (25%). The mean number of assessments completed per participant was 2.26, SD=2.01. 18% of baseline participants provided a 3-month assessment, and 5% provided a 6-month assessment (see Figure 1 for patient flow diagram).

**Table 1.** Sample Characteristics.

| **Baseline and Time-Invariant Variables** | **N** | **Mean** | **SD** |
| --- | --- | --- | --- |
| Age | 1806 | 42.12 | 11.60 |
| Frequency of Ketamine Doses Administered | 1806 | 4.04 | 2.70 |
| Frequency of Integration Sessions Delivered | 1711 | 3.21 | 2.03 |
|  | **N** | **Frequency** | **Percent** |
| Gender |  |  |  |
| Female | 1806 | 948 | 52.49 |
| Male | 1806 | 850 | 47.07 |
| Non-Binary | 1806 | 8 | 0.44 |
| Primary Diagnosis |  |  |  |
| Anxiety | 1745 | 488 | 27.97 |
| Bipolar Disorder | 1745 | 16 | 0.92 |
| Depression | 1745 | 418 | 23.95 |
| Eating Disorder | 1745 | 27 | 1.55 |
| Obsessive-Compulsive Disorder | 1745 | 47 | 2.69 |
| PTSD | 1745 | 442 | 25.33 |
| Substance Abuse | 1745 | 85 | 4.87 |
| Other | 1745 | 222 | 12.72 |
| **Caseness at Baseline** |  |  |  |
| T0 PHQ ≥ 15 | 1805 | 821 | 45.48 |
| T0 GAD ≥ 10 | 1802 | 1137 | 63.10 |
| T0 PCL ≥ 14 | 1797 | 1364 | 75.90 |
| **Time-Dependent Variables** | **N** | **Mean** | **SD** |
| T0 PHQ | 1805 | 13.60 | 6.51 |
| T1 PHQ | 331 | 10.26 | 6.78 |
| T3 PHQ | 316 | 8.29 | 6.02 |
| T6 PHQ | 92 | 8.92 | 6.06 |
| T0 GAD | 1802 | 11.86 | 5.60 |
| T1 GAD | 334 | 8.79 | 5.84 |
| T3 GAD | 306 | 6.73 | 5.03 |
| T6 GAD | 72 | 8.39 | 5.84 |
| T0 PCL | 1797 | 17.98 | 5.63 |
| T1 PCL | 328 | 15.30 | 5.49 |
| T3 PCL | 300 | 13.88 | 4.98 |
| T6 PCL | 69 | 15.22 | 5.48 |

T0 = baseline; T1 = 1-month assessment; T3 = 3-month assessment; T6 = 6-month assessment.

**Figure 1.**
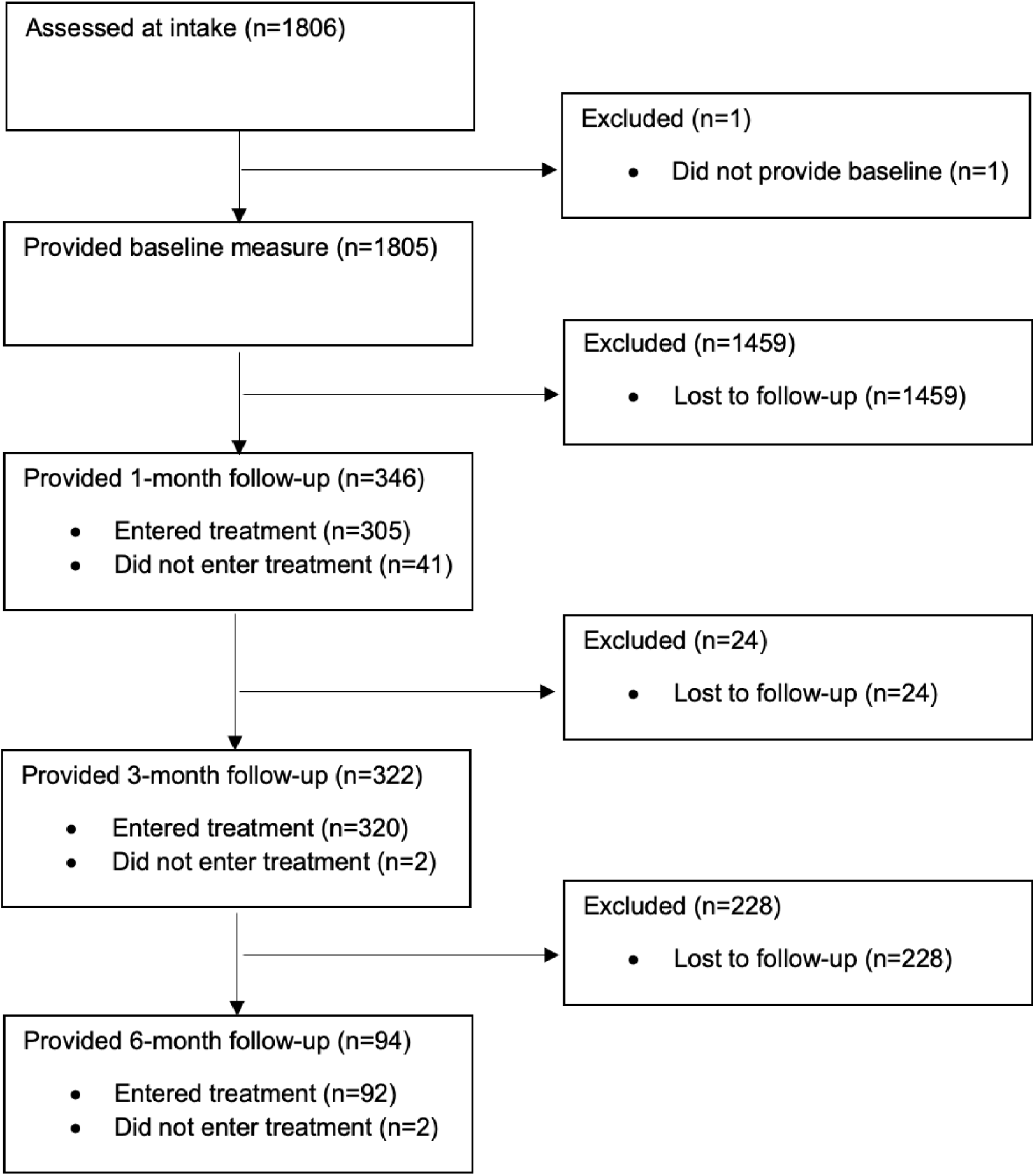
Patient Flow Diagram.

The mean number of doses was 4, SD=3, with 12% (210/1806) receiving more than 6 doses and 24% (440/1806) receiving 1 dose. Ketamine was administered sublingually to 32% (579/1806) of the sample and intramuscularly to 68% (1227/1806). The mean number of (psychotherapeutic) integration sessions was 3, SD=2, with 20% (365/1806) receiving more than 4 sessions.

### Lost to Follow-up Analyses

Lost to follow-up refers to individuals who did not provide a follow-up assessment and is separate from treatment status. Lost to follow-up was associated with less pretreatment psychological distress. Compared to those who provided a 3-month assessment, those who did not had lower baseline scores on PHQ, M_Diff_=−1.83, CI_.95_ (−1.09, −2.58), p<.0001, d=0.28, and PCL, M_Diff_=−1.56, CI_.95_ (−0.86, −2.25), p<.0001, d=0.28. Compared to those who provided a 6-month assessment, those who did not had lower baseline scores on GAD, M_Diff_=−1.58, CI_.95_ (−0.26, −2.90), p=0.02, d=0.28, and PCL, M_Diff_=−1.86, CI_.95_ (−0.51, −3.22), p=0.007, d=0.33. Overall, the frequency of missing assessments had a tendency to be negatively correlated with baseline scores on the PHQ, r=−0.13, p<.0001; the GAD, r=−0.06, p=0.02; and the PCL, r=−0.10, p<.0001.

### Primary and Secondary Outcomes

The primary outcomes were changes at 3 months from baseline, and secondary outcomes were changes at 1 and 6 months from baseline. There was a significant reduction on the PHQ at 1 month, d=0.50, which was amplified at 3 months, d=0.85, and remained detectable at 6 months, d=0.73. There was a reduction on the GAD at 1 month, d=0.47, which was amplified at 3 months, d=0.86, and remained detectable at 6 months, d=0.73. There was a reduction on the PCL at 1 month, d=0.38, which was amplified at 3 months, d=0.75, and remained detectable at 6 months, d=0.61. See Table 2 for details.

**Table 2.** Predicted Mean Differences from Linear Mixed Model Analyses of Available Cases and Multiple Imputation.

| Outcome | M1<br>Time<br>point | M2<br>Time<br>point | M1 | M2 | diff<sub>M1-M2</sub> | Lower<br>CL<sub>-.95</sub> | Upper<br>CL<sub>.95</sub> | d | Imputed<br>diff<sub>M1-M2</sub> | ImputedLower<br>CL<sub>-.95</sub> | Imputed<br>Upper<br>CL<sub>.95</sub> | Imputed<br>d |
| --- | --- | --- | --- | --- | --- | --- | --- | --- | --- | --- | --- | --- |
| PHQ | T1 | T0 | 10.38 | 13.62 | -3.23*** | -3.57 | -2.90 | 0.50 | -3.88*** | -4.49 | -3.28 | 0.60 |
|  | T3 | T0 | 8.06 | 13.62 | -5.56*** | -5.96 | -5.15 | 0.85 | -5.96*** | -6.57 | -5.34 | 0.92 |
|  | T6 | T0 | 8.88 | 13.62 | -4.74*** | -5.68 | -3.80 | 0.73 | -4.79*** | -5.83 | -3.75 | 0.74 |
| GAD | T1 | T0 | 9.24 | 11.89 | -2.65*** | -2.95 | -2.34 | 0.47 | -3.16*** | -3.71 | -2.61 | 0.56 |
|  | T3 | T0 | 7.09 | 11.89 | -4.80*** | -5.17 | -4.44 | 0.86 | -5.27*** | -5.80 | -4.74 | 0.94 |
|  | T6 | T0 | 7.78 | 11.89 | -4.10*** | -5.02 | -3.18 | 0.73 | -4.04*** | -5.24 | -2.84 | 0.72 |
| PCL | T1 | T0 | 15.85 | 17.99 | -2.14*** | -2.44 | -1.83 | 0.38 | -2.72*** | -3.26 | -2.18 | 0.48 |
|  | T3 | T0 | 13.75 | 17.99 | -4.24*** | -4.60 | -3.87 | 0.75 | -4.53*** | -5.08 | -3.98 | 0.80 |
|  | T6 | T0 | 14.55 | 17.99 | -3.44*** | -4.38 | -2.50 | 0.61 | -3.57*** | -4.71 | -2.42 | 0.63 |

T0 = baseline; T1 = 1-month assessment; T3 = 3-month assessment; T6 = 6-month assessment.
\*\*\*p<.0001.

### Dose and Other Predictive Factors

We entered dose as a predictive factor, controlling for age, gender, and site. Dose referred to the number of doses administered at the time of assessment and was a within-subjects, time-varying factor. There was a significant effect of dose on PHQ, b=−1.20, se=0.061, p<.0001, on GAD, b=−0.97, se=0.055, p<.0001, and on PCL, b=−0.92, se=0.055, p<.0001. These coefficients indicate that per administered dose, there was a reduction of 1.20 points on the PHQ and approximately 1 point reductions on the GAD and PCL.

There were main effects of site, age, and gender. There were medium to large site differences due to the requirement for clinician treatment referral in Canada. Overall, Canadian sites treated cases with more intense symptomatology than American sites (which permitted self-referral): d=0.80, M_Diff_=5.19, CI_.95_ (4.19, 6.20) on PHQ; d=0.43, M_Diff_=2.43, CI_.95_ (1.58, 3.28) on GAD; d=0.58, M_Diff_=3.27, CI_.95_ (2.41, 4.14) on PCL; all p’s<.0001. There were some small age effects. Compared to younger clients (estimated at 30 years or mean age – 1SD), older clients (estimated at 54 years or mean age + 1SD) had lower overall PHQ scores, d=0.23, M_Diff_=−1.53, CI_.95_ (−2.09, −0.98), p<.0001, lower overall GAD scores, d=0.31, M_Diff_=−1.73, CI_.95_ (−2.20, −1.25), p<.0001, and lower overall PCL scores, d=0.14, M_Diff_=−0.78, CI_.95_ (−1.27, −0.30), p=0.002. There was one marginally significant gender difference. Compared to men, women had a slight tendency to report higher overall PHQ scores, d=0.08, M_Diff_=0.54, CI_.95_ (−0.005, 1.08), p=0.05.

### Caseness

There were statistically significant reductions in the proportions of depressed, anxious, and PTSD cases as identified by scoring above or equal to clinical cut-off values on each outcome measure (see Table 3). Case reductions showed similar patterns of change across outcomes as in the growth curve models, with the largest case reductions at the primary 3-month endpoint.

**Table 3.**
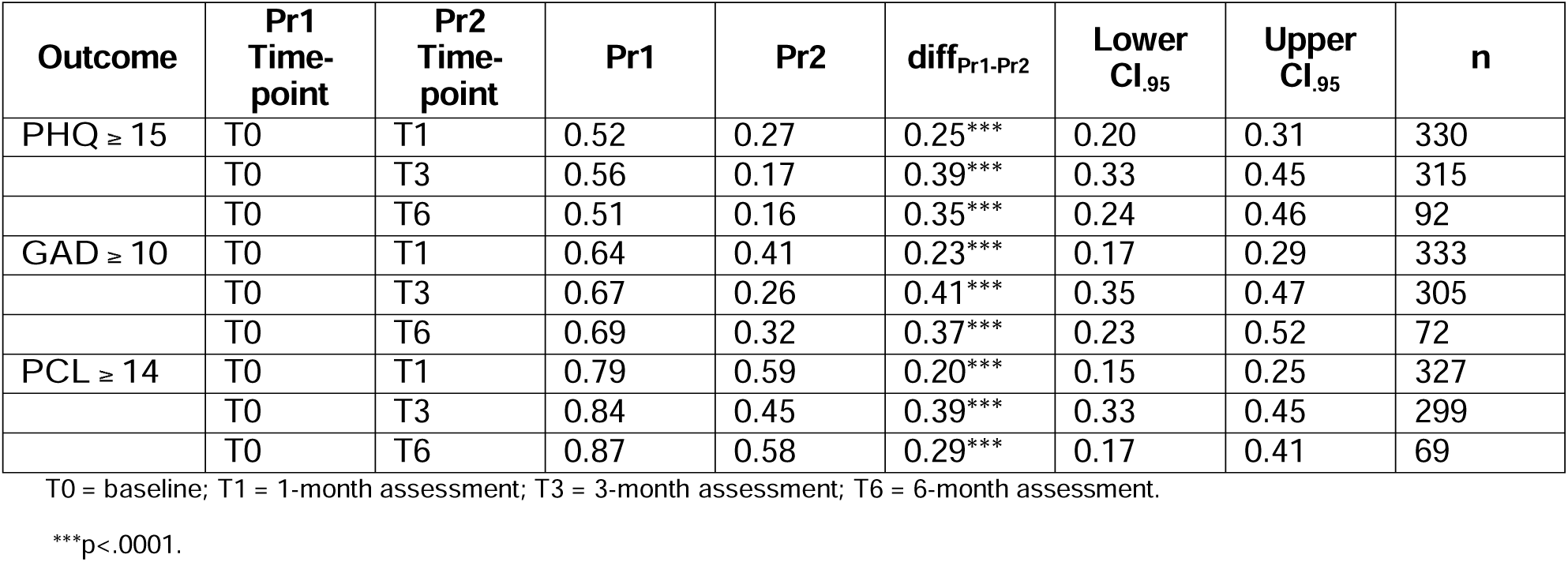
Proportions scoring above cut-off by outcome and timepoint.

| **Outcome** | **Pr1**<br>**Time-**<br>**point** | **Pr2**<br>**Time-**<br>**point** | **Pr1** | **Pr2** | **diff<sub>Pr1-Pr2</sub>** | **Lower**<br>**CI<sub>.95</sub>** | **Upper**<br>**CI<sub>.95</sub>** | **n** |
| --- | --- | --- | --- | --- | --- | --- | --- | --- |
| PHQ ≥ 15 | T0 | T1 | 0.52 | 0.27 | 0.25*** | 0.20 | 0.31 | 330 |
|  | T0 | T3 | 0.56 | 0.17 | 0.39*** | 0.33 | 0.45 | 315 |
|  | T0 | T6 | 0.51 | 0.16 | 0.35*** | 0.24 | 0.46 | 92 |
| GAD ≥ 10 | T0 | T1 | 0.64 | 0.41 | 0.23*** | 0.17 | 0.29 | 333 |
|  | T0 | T3 | 0.67 | 0.26 | 0.41*** | 0.35 | 0.47 | 305 |
|  | T0 | T6 | 0.69 | 0.32 | 0.37*** | 0.23 | 0.52 | 72 |
| PCL ≥ 14 | T0 | T1 | 0.79 | 0.59 | 0.20*** | 0.15 | 0.25 | 327 |
|  | T0 | T3 | 0.84 | 0.45 | 0.39*** | 0.33 | 0.45 | 299 |
|  | T0 | T6 | 0.87 | 0.58 | 0.29*** | 0.17 | 0.41 | 69 |

T0 = baseline; T1 = 1-month assessment; T3 = 3-month assessment; T6 = 6-month assessment.
\*\*\*p<.0001.

In the 315 patients who provided a 3-month PHQ assessment, the depression case rate dropped from 56% at baseline to 17% at 3 months. In the 305 patients who provided a 3-month GAD assessment, the anxiety case rate dropped from 67% at baseline to 26% at 3 months. In the 299 patients who provided a 3-month PCL assessment, the PTSD case rate dropped from 84% at baseline to 45% at 3 months.

In the 92 patients who provided a 6-month PHQ assessment, the depression case rate dropped from 51% at baseline to 16% at 6 months. In the 72 patients who provided a 6-month GAD assessment, the anxiety case rate dropped from 69% at baseline to 32% at 6 months. In the 69 patients who provided a 6-month PCL assessment, the PTSD case rate dropped from 87% at baseline to 58% at 6 months.

### Treatment Responders

See Table 4. In the 315 patients who provided a 3-month PHQ assessment, 75% reported a 3-point MCID compared to baseline. In the 305 patients who provided a 3-month GAD assessment, 68% reported a 3-point MCID compared to baseline. In the 299 patients who provided a 3-month PCL assessment, 50% reported a 5-point MCID compared to baseline.

**Table 4.** Proportions reporting MCIDs at endpoint compared to baseline.

| **Outcome** | **Endpoint** | **Pr** | **Lower CI<sub>.95</sub>** | **Upper CI<sub>.95</sub>** | **n** |
| --- | --- | --- | --- | --- | --- |
| PHQ | T1 | 0.53*** | 0.47 | 0.58 | 330 |
|  | T3 | 0.75*** | 0.70 | 0.79 | 315 |
|  | T6 | 0.70*** | 0.60 | 0.79 | 92 |
| GAD | T1 | 0.50*** | 0.44 | 0.55 | 333 |
|  | T3 | 0.68*** | 0.62 | 0.73 | 305 |
|  | T6 | 0.65*** | 0.54 | 0.76 | 72 |
| PCL | T1 | 0.32*** | 0.26 | 0.37 | 327 |
|  | T3 | 0.50*** | 0.44 | 0.56 | 299 |
|  | T6 | 0.48*** | 0.36 | 0.60 | 69 |

Note. MCID = 3 points for PHQ and GAD. MCID = 5 points for PCL.
T1 = 1-month assessment; T3 = 3-month assessment; T6 = 6-month assessment.
\*\*\*p<.0001.

In the 92 patients who provided a 6-month PHQ assessment, 70% reported a 3-point MCID compared to baseline. In the 72 patients who provided a 6-month GAD assessment, 65% reported a 3-point MCID compared to baseline. In the 69 patients who provided a 6-month PCL assessment, 48% reported a 5-point MCID compared to baseline.

### Simulations

We multiply imputed 1000 datasets per outcome measure in which missing values were filled in with simulated values, achieving relative efficiencies of 0.999. This method allows us to evaluate what would have happened if individuals who were lost to follow-up were retained in the data analysis and is meant to mitigate the poor retention rate. For each imputation, we specified linear mixed models to directly estimate mean differences at each endpoint from baseline. These parameters were combined across imputations to achieve best estimates of mean differences (see Table 2). There were no apparent differences between imputed estimates and those based on the analysis of available cases only. Many imputed estimates indicated a slightly larger effect than initially detected.

## Discussion

This single-arm effectiveness trial examined the largest dataset to date of long term outcomes in clients who received Ketamine-Assisted Psychotherapy. KAP aimed to alleviate symptoms of depression, anxiety, and post traumatic stress. We found evidence of large to moderately large treatment effects at 3 and 6 months, d’s ranging from 0.61-0.86 and corresponding to 5-6 point reductions on the PHQ, 4-5 point reductions on GAD, and 3-4 point reductions on PCL. There was a consistent pattern of change across measures characterized by reductions in psychological distress that were most apparent at 3 months. There were approximately 40% reductions in caseness for depression, anxiety, and PTSD at this primary endpoint, and treatment responders ranged from 50-75%. This clinical benefit was sustained at 6 months.

Gender and age differences in outcomes were small to minimal, indicating the broad suitability of KAP. Over 90% of individuals entered treatment within 3 months of intake, and 76% completed more than 1 ketamine dose. Each ketamine dose was associated with 1-point reductions on the PHQ, GAD, and PCL, in support of the current program, which recommends 4-6 ketamine sessions. This can be anticipated to produce an estimated 4-6 point reduction across outcome measures over time, larger than the MCID thresholds of 3-5 points for indicating treatment response. However, we caution that the effects of dose must be interpreted within the context of KAP, which involved integration sessions that occurred in lock step with ketamine dosing to activate and reinforce the psychological gains from this experience. Occasional side effects included nausea, vomiting, and increases in blood pressure, not requiring medical intervention nor requiring patients to be taken to a hospital. Findings may be most generalizable to populations with treatment-resistant depression, anxiety, or PTSD eligible for KAP treatment according to Fieldtrip Health guidelines. Given the lack of novel treatment options in the behavioral health space, KAP represents a viable alternative that should be more widely considered.

## Limitations

The main limitation of this study is the substantial attrition during the 6 month follow-up period. The importance of completing the follow-up assessments was not made clear to patients and increased efforts are needed to improve survey completion rates moving forward. The treatment effects demonstrated here are based on those who provided follow-up data. These individuals tended to have more pretreatment distress and may be considered a clinical priority. The estimates of treatment effect at 6 months are less reliable than at 3 months. To the extent possible, we examined these issues using multiple imputation of missing data. The estimates of treatment effect from these simulation analyses were stable and consistent with the analysis of only available cases.

Although promising, a future prospective clinical trial with improved data collection is needed. Other limitations include the retrospective nature of the study and lack of a randomised control group. Further study is also needed to clarify the indications and contraindications of KAP for patient subpopulations and the tailoring of treatment to further improve upon individual outcomes.

## Conclusion

This large retrospective single-arm trial found that Ketamine-Assisted Psychotherapy is an effective treatment achieving sustained and clinically meaningful reductions in depression, anxiety, and post traumatic stress for up to half a year.

## Data Availability

There is no plan to release the data outside of reports of summative indices, but requests for access will be considered. 

## ARTICLE INFORMATION

## Author Contributions

Drs Yermus and Lo had full access to all of the data in the study and take responsibility for the integrity of the data and the accuracy of the data analysis.

*Concept and design*: Yermus, Verbora, Lo.

*Acquisition, analysis, or interpretation of data:* All authors.

*Drafting of the manuscript:* All authors.

*Critical revision of the manuscript for important intellectual content:* All authors.

*Statistical analysis:* Setlur, Bottos, Yermus, Lo.

*Obtained funding:* Yermus, Lo.

*Administrative, technical, or material support:* Setlur, Zaer, Bottos.

*Supervision:* Yermus, Lo.

### Funding/Support

This publication of this study was supported by the Field Trip Health & Wellness Research Grant.

### Data Sharing Statement

There is no plan to share data, although requests may be made for access.

### Trial Protocol

Trial protocol will be available at clinicaltrials.gov after the results are published.

